# Seroprevalence of SARS-CoV-2 antibodies in children - A prospective multicentre cohort study

**DOI:** 10.1101/2020.08.31.20183095

**Authors:** Thomas Waterfield, Chris Watson, Rebecca Moore, Kathryn Ferris, Claire Tonry, Alison P Watt, Claire McGinn, Steven Foster, Jennifer Evans, Mark D Lyttle, Shazaad Ahmad, Shamez Ladhani, Michael Corr, Lisa McFetridge, Hannah Mitchell, Kevin Brown, Gayatri Amirthalingam, Julie-Ann Maney, Sharon Christie

**Author notes:** Corresponding author – Thomas Waterfield – Centre For Experimental Medicine, Wellcome Wolfson Institute of Experimental Medicine, Queen’s University Belfast, Belfast, UK.

## Abstract

**Background:** Studies based on molecular testing of oral/nasal swabs underestimate SARS-CoV-2 infection due to issues with test sensitivity and timing of testing. The objective of this study was to report the presence of SARS-CoV-2 antibodies, consistent with previous infection, and to report the symptomatology of infection in children.

**Design:** This multicentre observational cohort study, conducted between 16^th^ April – 3^rd^ July 2020 at 5 UK sites, aimed to recruit 900 children aged 2 to 15 years of age. Participants provided blood samples for SARS-CoV-2 antibody testing and data were gathered regarding unwell contacts and symptoms.

**Results:** 1007 participants were enrolled, and 992 were included in the final analysis. The median age of participants was 10·1 years. There were 68 (6.9%) participants with positive SARS-CoV-2 antibody tests indicative of previous SARS-CoV-2 infection. Of these, 34/68 (50%) reported no symptoms. The presence of antibodies and the mean antibody titre was not influenced by age. Following multivariate analysis 4 independent variables were identified as significantly associated with SARS-CoV-2 infection. These were: known infected household contact; fatigue; gastrointestinal symptoms; and changes in sense of smell or taste.

**Discussion:** In this study children demonstrated similar antibody titres in response to SARS-CoV-2 irrespective of age. The symptoms of SARS-CoV-2 infection in children were subtle but of those reported, fatigue, gastrointestinal symptoms and changes in sense of smell or taste were most strongly associated with antibody positivity.

**Registration:** This study was registered at https://www.clinicaltrials.gov (trial registration: NCT04347408) on the 15/04/2020.

## Introduction

During the first wave of the SARS-CoV-2 pandemic in England, children accounted for just 1% of confirmed infections,(1) had a milder clinical course, and had a much lower mortality than adults (1-4), a pattern similar to other international settings (3,4). The reasons for this are unknown, but various hypothesises exist. Public health measures, such as school closures, may have minimised children’s exposure to SARS-CoV-2. It is also possible that children have a different immune response to the virus for example the reduced expression of the ACE2 gene, the host receptor for SAR-CoV-2 virus in airway cells (5-7).

Despite existing data, it is impossible to state accurately what proportion of children were infected with SARS-CoV-2 in the UK. Studies based on molecular testing of oral/nasal swabs with real-time reverse transcription polymerase chain reaction (RT-qPCR) underestimate infection due to issues with test sensitivity, timing of testing and non-testing of asymptomatic individuals (8). A potentially more reliable method is to test for specific antibodies. Existing antibody tests typically detect immunoglobulin G (IgG or Total antibody) to either the nucleocapsid or spike proteins of the virus (9). Antibody testing has greater potential than RT-qPCR to detect previous asymptomatic/mildly symptomatic infection, and is not dependent on coinciding with active infection. Current best seroprevalence estimates from adults in the UK indicate that approximately 6.2% have antibodies consistent with previous SARS-CoV-2 infection (10). These findings are similar to other international seroprevalence studies (11-13). Currently there are no published data reporting the current seroprevalence of SARS-CoV-2 antibodies in UK children.

It is unclear what proportion of children are asymptomatic and which symptoms are most associated with paediatric SARS-CoV-2 infection. Estimates based on RT-qPCR testing of oral/nasal swabs suggest that cough or fever are the most common symptoms (14-19). However, these studies focus on symptomatic cohorts, introducing selection bias (14-19), which leads to underestimation of the asymptomatic proportion.

The objective of this study was to report the presence, and titres, of SARS-CoV-2 antibodies in healthy children of healthcare workers across the UK and to report the symptomatology of infection including the asymptomatic rate.

## Methods

### Study Design

This multicentre observational prospective cohort study was designed to determine the seroprevalence of SARS-CoV-2 antibodies in healthy children, and report the symptomatology of infection. This study has been written in conjunction with the Strengthening the Reporting of Observational Studies in Epidemiology (STROBE) guidelines (20). The study protocol has undergone external peer review and is available as an open access publication (21).

### Setting

Participants were recruited from 5 UK centres, in the 4 regions of the UK, between 16^th^ April 2020 and 3^rd^ July 2020. The sites included tertiary NHS hospitals (Belfast, Cardiff, Manchester, and Glasgow) and a Public Health England site (London).

### Participants

Children of healthcare workers, aged between 2 and 15 years at the time of recruitment, were eligible to participate. A “healthcare worker” was defined as a National Health Service (NHS) employee. Healthcare workers were categorised according to role, including whether that role involved patient facing activities. Approximately 150 non-patient facing staff were included to provide a comparison group, and to improve the generalisability of the results. Participants were identified at each participating NHS organisation using internal intranet advertisements and email circulars. Children were excluded if they were receiving antibiotics, had been admitted to hospital within the last 7 days, were receiving oral immunosuppressive treatment, or if ever diagnosed with a malignancy.

### Informed consent

Informed consent was obtained, and assent given by children where possible. Participants were free to decline/withdraw consent at any time without providing a reason and without being subject to any resulting detriment.

### Assessments and procedures

All children underwent phlebotomy performed by experienced paediatric medical and nursing professionals. Serum and/or plasma were tested for antibodies to SARS-CoV-2, in UKAS accredited laboratories using the following assays, which have been validated for use (22-24):

- Nucleocapsid assays – (Abbott Architect® SARS-CoV-2 IgG and Roche Elecsys® Anti-SARS-CoV-2 Total Antibody)
- Spike protein assays – (DiaSorin LIAISON® SARS CoV-2 S1/S2 IgG assay)

The Abbott, Roche and DiaSorin assays are highly specific for SARS-CoV-2 antibodies, using the manufacturer’s suggested cut-offs, with specificities of 1.00 (95% CI 0.98 to 1.00), 1.00 (95% CI 0.99 to 1.00) and 0.98 (95% CI 0.96 to 0.99) respectively (22-24). They do however have lower sensitivities at 0.94 (95% CI 0.86 to 0.98), 0.84 (95% CI 0.75 to 0.91) and 0.64 (95% CI 0.54 to 0.73) respectively (22-24). A summary of the tests used is provided in Table 1.

**Table 1:** Summary of antibody tests used

| *Name of assay* | *Target* | *Units* | *Cut-Off* |
| --- | --- | --- | --- |
| *Abbott Architect® SARS-CoV-2 IgG* | Nucleocapsid | Calculated index S/C | 1.4 S/C |
| *Roche Elecsys® Anti-SARS-CoV-2* | Nucleocapsid | Cut-off index COI | 1.0 COI |
| *DiaSorin LIAISON® SARS CoV-2 S1/S2 IgG assay* | Spike protein | Arbitrary units AU/ml | 15.0 AU/ml |

Study data were collected on a case report form (CRF) using REDCap (Research Electronic Data Capture) electronic data capture tools (25). Participants and their parents provided information at enrollment relating to age, sex, previous health and potential predictors of SARS-CoV-2 infection including; known contact with individuals with COVID-19, contact with individuals who have been symptomatic and/or self-isolating and results of any diagnostic testing such as RT-qPCR testing/antibody testing. To minimise recall bias, data relating to exposures and illness episodes were collected blinded to antibody testing results.

### Outcome Measures

- Presence of antibodies (IgG/Total antibody) to SARS-CoV-2 in serum or plasma reported as titres.
- Previous SARS-CoV-2 infection defined as a positive antibody test using the manufacturer’s advised positivity cut-off.

### Sample Size Justification

The study was powered to detect a change in seroprevalence of SARS-CoV-2 antibodies at 3 time-points (enrollment, and 2 and 6 months following enrollment). To achieve this, 675 participants were required (assuming alpha of, 0.05 and beta of 0.2). Allowing for 30% dropout rate, we aimed to recruit 900 participants from 5 sites.

### Statistical analysis plan

Variables including sex, age, parent role, symptomatology, household contacts, and SARS-CoV-2 antibody prevalence were analysed using descriptive statistics (number and proportion for discrete variables, median and interquartile range for continuous variables). Seroprevalence rates between sites were compared using Fisher’s exact test and antibody titres were correlated with age using the Kendall’s rank correlation test and mean titres were compared between symptomatic and asymptomatic participants using the Wilcoxon rank sum test.

Variables associated with SARS-CoV-2 infection were analysed in a stepwise approach. Initially all possible variables were assessed using univariate analysis with Fisher’s exact testing of categorical data, and the Mann-Whitney U test for continuous data (continuous data were skewed). All variables with a statistically significant association with SARS-CoV-2 infection (p<0.20) were included in a weighted binary multivariate logistic regression model. A liberal level of significance (p<0.20) was chosen to avoid falsely excluding a significant variable based on univariate analysis alone. Participants with incomplete CRFs were excluded from univariate and multivariate analysis. Analysis was conducted in R (R Core Team, 2014). *Patient and Public Involvement (PPI)*

A PPI group comprising parents and children was convened. The PPI group met virtually and via socially distanced meetings. The group have contributed to the design of the study through online surveys and video discussions. They have also contributed to media interviews on national television and the lead young person has co-authored a manuscript outlining their experience of taking part in the study (26).

### Office for Research Ethics Committees (OREC) and local Research Governance

Ethical approval was obtained from the London – Chelsea Research Ethics Committee (REC Reference – 20/HRA/1731) and the Belfast Health & Social Care Trust Research Governance (Reference 19147TW-SW).

### Study Registration

This study was registered at https://www.clinicaltrials.gov (trial registration: NCT04347408) on the 15/04/2020 (last updated 27/05/20). At the time of registration no patients had been recruited to the study which opened on the 16/04/20. The end of the study will be the last study visit.

## Findings

In total, 1042 potential participants were screened for inclusion, of whom 35 were excluded; 18 were outside the specified age range, 1 met specific exclusion criteria, and 16 declined consent. The remaining 1007 children were enrolled, of which 15 were excluded from analysis due to unsuccessful phlebotomy; 992 were included in the final analysis (Figure 1). The recruitment by site can be visualised in Table 2. In the analysis cohort 962/992 (97%) had complete CRFs and 30/992 (3%) had partially complete CRFs.

**Table 2:**
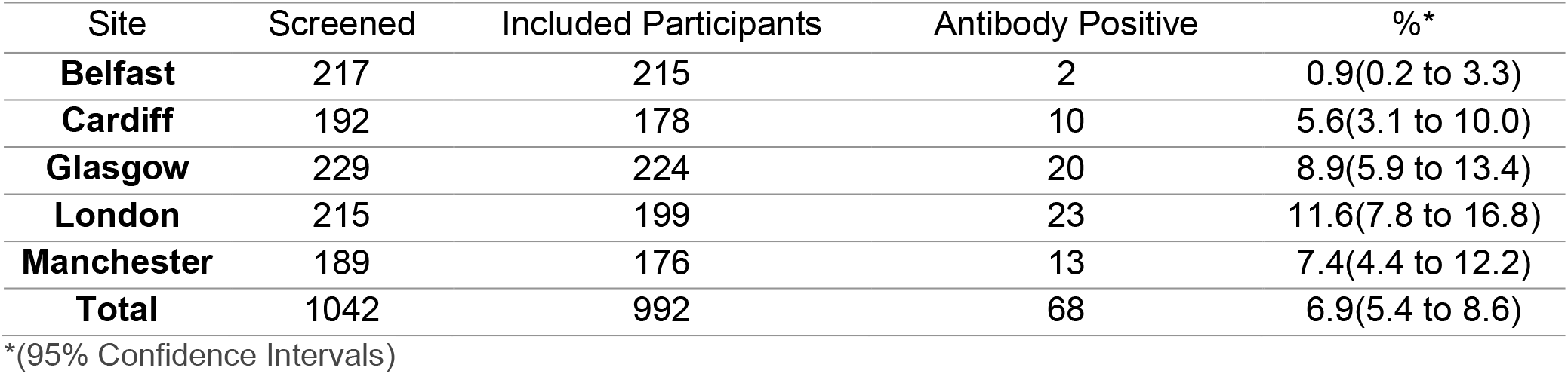
Recruitment summary and seroprevalence by site (n and (%) unless otherwise stated)

**Figure 1:**
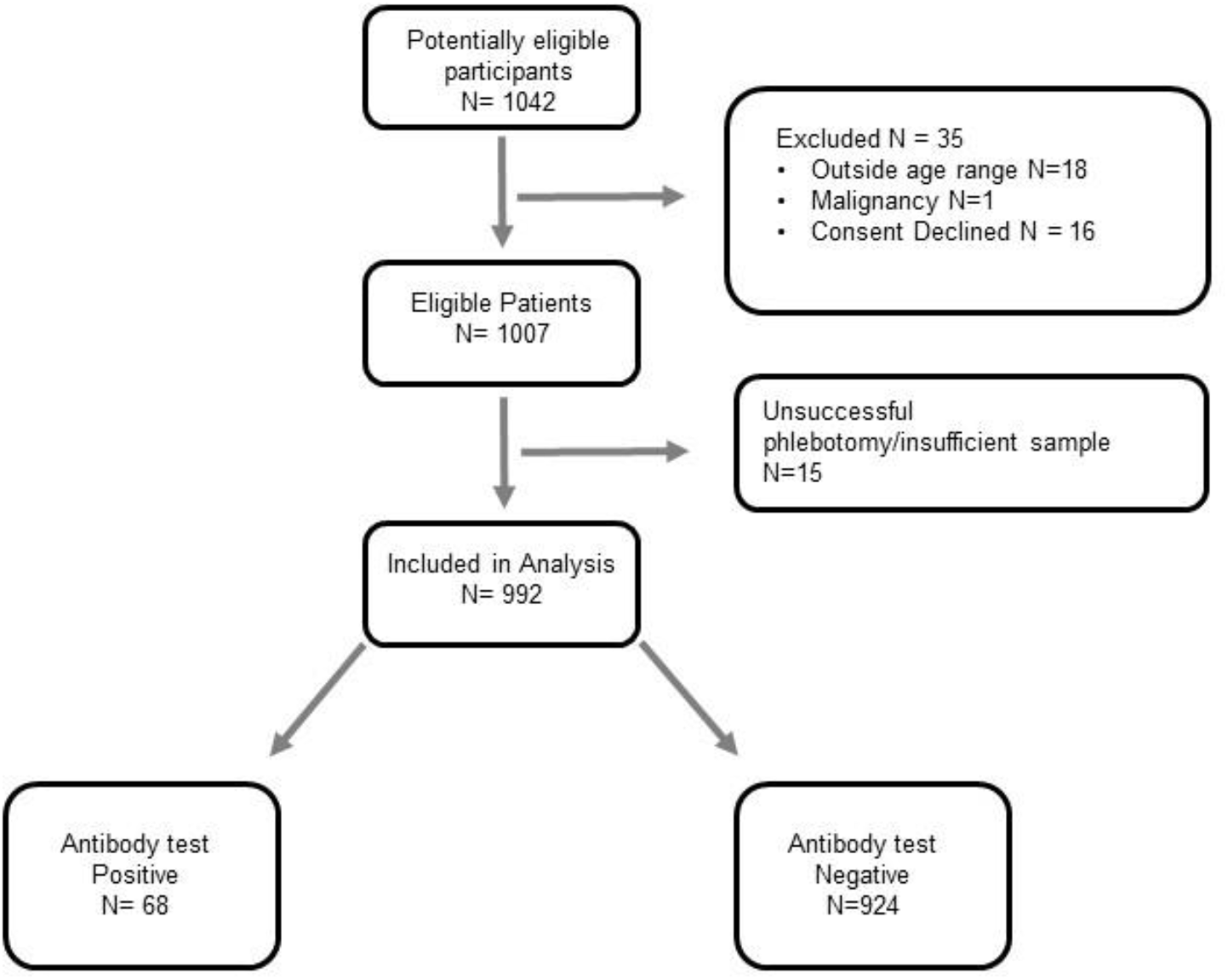
Flow of patients through the study

The median age of participants was 10·1 years (range 2.03 to 15.99 years), with 484 (49%) aged under 10 years; 509 (51%) were male. There were 68/992 participants with positive SARS-CoV-2 antibodies, giving a seroprevalence of 6.9% (95% CI 5.4 to 8.6, n=992). Of those with positive SARS-CoV-2 antibody tests, 34/68 (50%) reported no symptoms. The most commonly reported symptoms of SARS-CoV-2 infection were fever 21/68 (31%), gastrointestinal symptoms (diarrhoea, vomiting and abdominal cramps) 13/68 (19%) and headache 12/68 (18%). The presence of fever, cough or changes in a sense of smell/taste were recorded in 26/68 (38%) of participants. No children within this cohort had severe disease requiring hospital admission. A summary of reported symptoms and their frequency can be seen in Table 3.

**Table 3:**
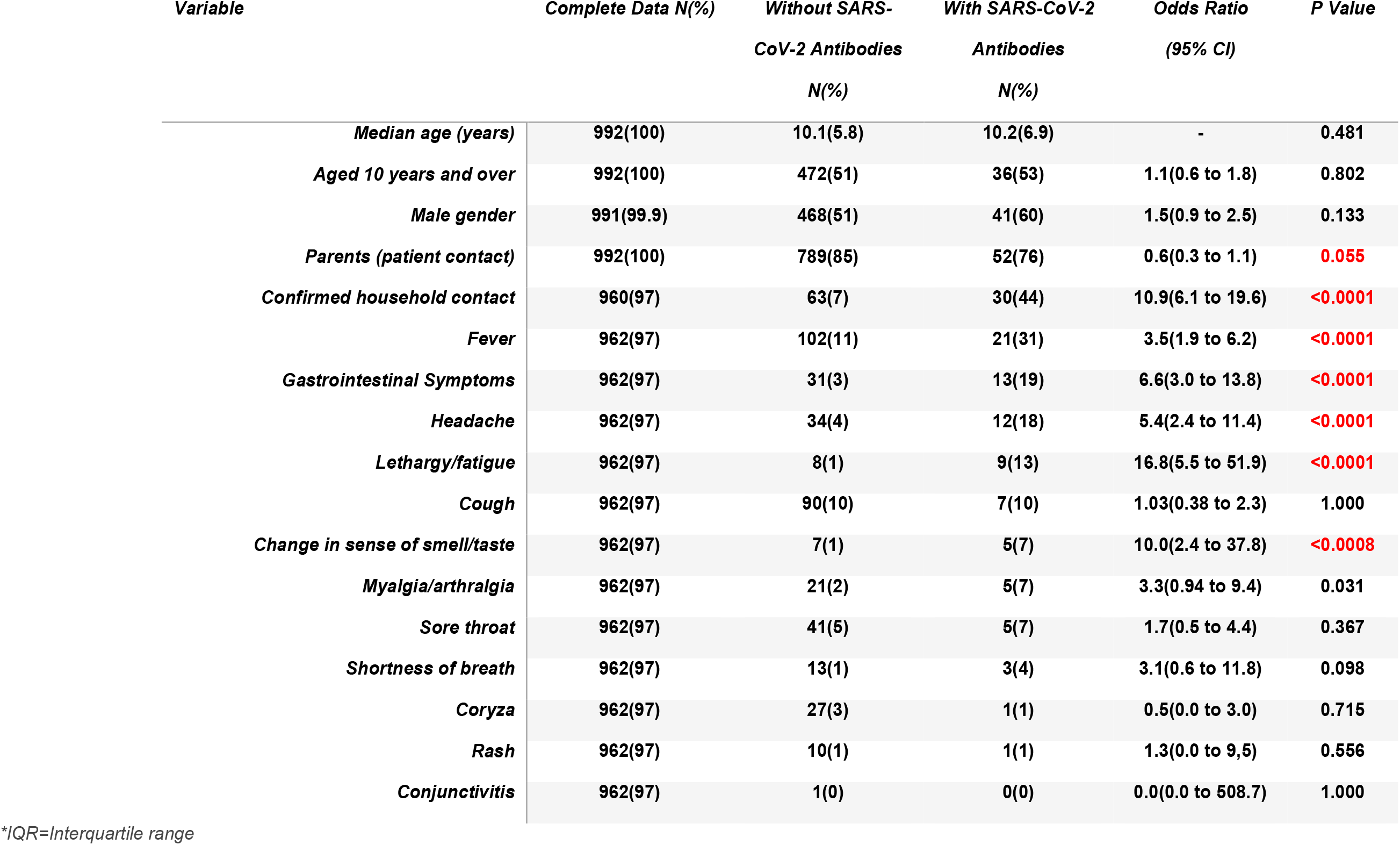
Univariate analysis of variables (Fisher’s Exact for categorical variables, Mann-Whitney U for continuous variables). Number and (%) with feature shown for categorical variables and median for continuous variables unless otherwise stated.

Seroprevalence of SARS-CoV-2 antibodies varied between sites. Belfast had significantly lower seroprevalence than all other sites at 0.9% (95% CI 0.2 to 3.3, n=215); p<0.0001, and in London seroprevalence was significantly higher than all other sites at 11.6% (95% CI 7.8 to 16.8 n=199); p=0.0069. The remaining 3 sites reported seroprevalence rates between 5.6% and 8.9%. The difference between these 3 sites were not significant (Table 2)..

The mean antibody titres, for those testing positive, were;

- 4.86 S/C (95%CI 4.28 to 5.45, n=58) for the Abbott Architect® SARS-CoV-2 IgG assay.
- 65.32 COI (95% CI 43.24 to 87.40, n=31) for the Roche Elecsys® Anti-SARS-CoV-2 Total Antibody assay.
- 64.17 AU/ml (95% CI 37.99 to 90.36, n=31) for the DiaSorin LIAISON® SARS CoV-2 S1/S2 IgG assay. T

There was no correlation between age and antibody titres (Figure 2). The results from the Abbott Architect® SARS-CoV-2 IgG assay indicated a small but significant difference in mean antibody titres between asymptomatic 4.3 S/C (95% CI 3.4 to 5.2) and symptomatic participants 5.5 S/C (95% CI 4.7 to 6.2); p=0.04. There was no significant difference in mean antibody titres for the Roche Elecsys® or DiaSorin LIAISON® assays when comparing symptomatic and asymptomatic participants (p =0.23 and 0.58 respectively) (Figure2).

**Figure 2:**
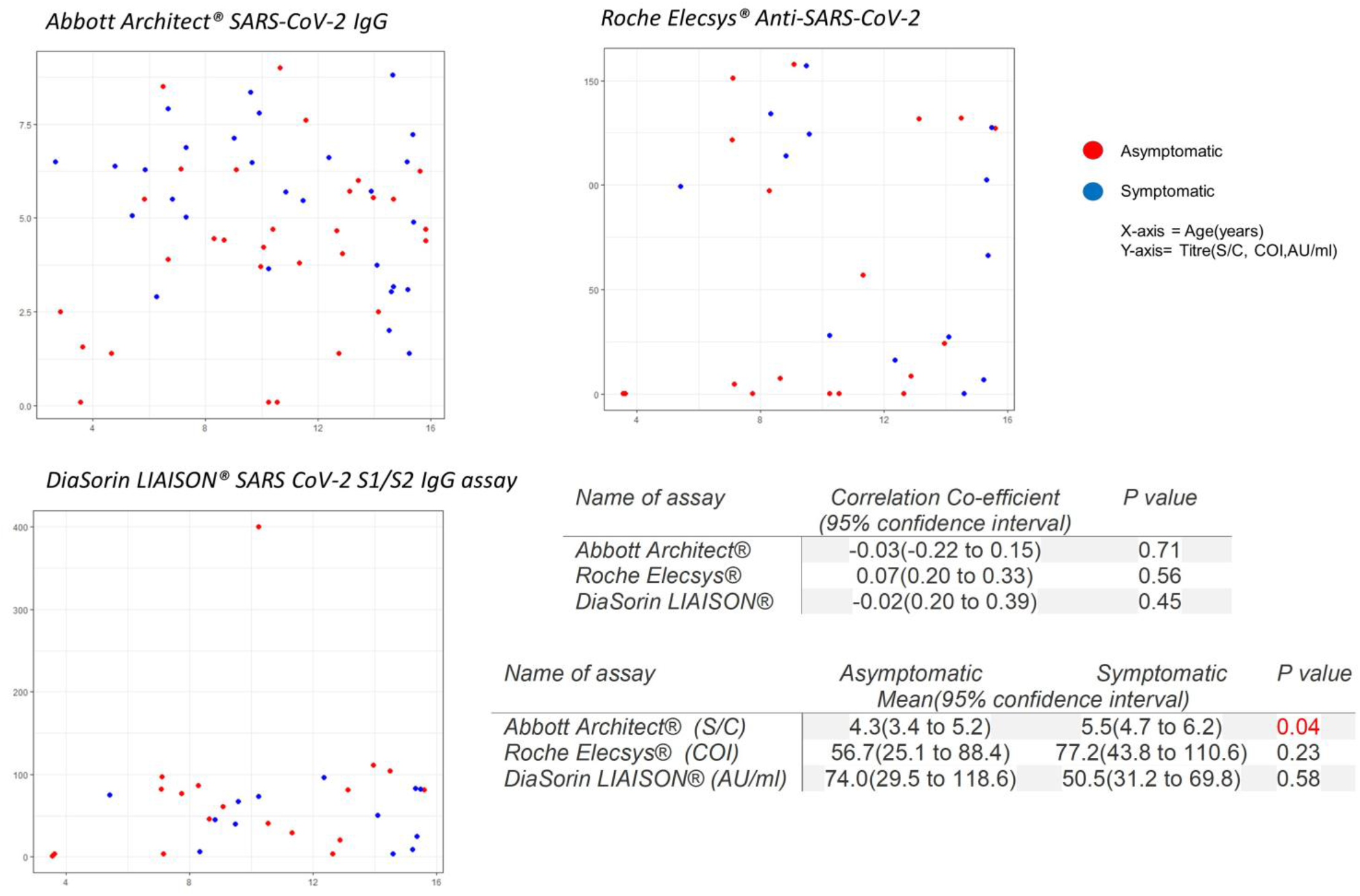
Scatter diagrams of age/symptoms and SARS-CoV-2 assay titre. Abbott Architect® reported in S/C, Roche Elecsys® reported in COI, DiaSorin LIAISON® reported in AU/ml.

The univariate analysis of individual variables associated with SARS-CoV-2 infection is shown in Table 3. In addition to clinical features, variables such as age, gender, the role of the parent (patient facing or not), and known household contacts were included. Age and gender were not associated with SARS-CoV-2 infection (Table 3). Parental role showed significant association in the univariate analysis, but this was no longer significant once corrected for site and other variables in the multivariate analysis. Contact with a household member with confirmed SARS-CoV-2 infection was significantly associated with SARS-CoV-2 infection in the participant in both the univariate and multivariate analyses (Table 3). The multivariate analysis identified 4 variables independently associated with the presence of SARS-CoV-2 antibodies: (i) known household contact with confirmed SARS-CoV-2 (p<0.0001), (ii) fatigue (p=0.001), (iii) gastrointestinal symptoms (p=0.0001), and (iv) changes in sense of smell or taste (p<0.0012).

## Interpretation

This observational study is one of the largest UK studies of paediatric SARS-CoV-2 antibody seroprevalence, and the only study to recruit from all regions of the UK. Following the first pandemic wave in the UK, 68/992 (6.9%) children of healthcare workers had evidence of prior infection with SARS-CoV-2. Whilst this is likely to be higher than the general population it is surprisingly similar to the seroprevalence reported by the ONS study of adults from England and Wales (6.2%) (10), and similar to international estimates (11-13). As expected there was marked geographical variation, with London reporting the highest infection rates (11.6%) and Belfast the lowest (0.9%) p<0.0001. These regional variations are consistent with published adult estimates of seroprevalence from the same time period (10).

In this study there was a near equal number of children under 10 years of age 32/68 (47%) and children over 10 years of age 36/68 (53%) developing antibodies consistent with previous SARS-CoV-2 infection. Age, as a categorical or continuous variable, was not a statistically significant factor in predicting the presence of antibodies, or the overall titres in children irrespective of the assay used (Figure 2). This is in contrast to several studies that have reported a lower seroprevalence in young children (under 10 years of age) and in elderly adults (over 65 years of age) following the first wave of the pandemic (11-13). This has led some authors to suggest that children are less susceptible to SARS-CoV-2 infection (27-30). The studies on which these assumptions are based have typically reported a binary antibody outcome (positive or negative) rather than absolute titres (11-13). It is possible that the lower seroprevalence reported thus far in younger children merely reflects the effect of social distancing measures on this group. This may go some way to explain why the over 65s also demonstrated lower seroprevalence in the same studies (27-30). In our cohort, children were more likely to be exposed to SARS-CoV-2 in the home due to fact that their parent(s) worked in healthcare. The findings from this study may therefore provide a greater insight into how younger children react when exposed to SARS-CoV-2. Further research is required to understand if younger children are really less susceptible to SARS-CoV-2.

Of the 68 participants with positive antibody tests, 34/68 (50%) reported no symptoms. The most commonly reported symptoms associated with SARS-CoV-2 infection were fever (21/68) 30% and gastrointestinal symptoms 13/68(19%). These symptoms, in addition to fatigue, and changes in sense of smell or taste, were independently associated with previous SARS-CoV- 2 infection based on the weighted binary multivariate regression modelling. These findings reflect a number of international studies (14-19). Current UK testing strategies directing testing only for those with fever, cough or changes in smell/taste would have identified 26/34 (76%) of symptomatic participants in this study (assuming 100% sensitivity and specificity of RT-qPCR swab testing). Adding gastrointestinal symptoms would have identified nearly all symptomatic cases 33/34(97%).

There is evidence from adult serological studies that those with severe illness develop a significantly greater antibody response than those with mild or asymptomatic disease (31-33). This has raised concerns that children, who typically have mild disease, may fail to develop a meaningful antibody response to SARS-CoV-2 infection. More recently, emerging adult data suggest that even asymptomatic adults are capable of mounting a potentially lasting and protective immune response (34-35). In our study antibody titres, measured using the Abbott Architect® SARS-CoV-2 IgG assay, were significantly higher in symptomatic children compared with asymptomatic children p=0.04. These findings were not replicated with either the Roche Elecsys® Anti-SARS-CoV-2 or DiaSorin LIAISON® SARS CoV-2 S1/S2 IgG assays. It therefore remains unclear to what extent the severity of symptoms in children influences the antibody response.

## Summary

This study demonstrates that approximately half of children are asymptomatic when infected with SARS-CoV-2 and that current UK testing strategies will fail to diagnose the majority of paediatric infections. This study also demonstrates that younger children were just as likely to become infected with SARS-CoV-2 as older children and that they are capable of mounting a similar antibody response.

## Strengths/Limitations

The strengths of this study are that is a large multicentre study including children from across the four nations of the UK. The findings are based on serological antibody testing rather than RT-qPCR testing of swabs and are therefore more likely to report the true asymptomatic rate and the true symptomatology of paediatric infection with SARS-CoV-2.

The limitations of this study are that all of the children in this study had only mild disease making comparison between severe and mild disease impossible. The children were also recruited from healthcare workers and the prevalence of antibodies is likely to be lower in the general population. The children of healthcare workers were chosen for a number of reasons. Firstly, the study was conducted during the lock-down phase of the pandemic response thereby making face-to-face discussions challenging due to a need to conform with social distancing rules. Healthcare workers were felt to be more likely to be able to understand the study and consent without the need for face-to-face discussions with members of the research team. Secondly, healthcare workers were at higher risk of exposure to SARS-CoV-2 and their children were more likely to be infected making a study of symptomatology more practical.

What is known about this topic?

- Children are relatively unaffected by the SARS-CoV-2 infection with very few requiring hospitalisation.
- Most children with SARS-CoV-2 infection are asymptomatic.
- Molecular testing of oral/nasal swabs underestimates SARS-CoV-2 infection.

What this study adds

- Gastrointestinal upset is a relatively common symptom of Covid-19 in children. Adding gastrointestinal upset to the list of symptoms triggering a test in children would improve case-finding.
- Asymptomatic and mildly symptomatic children are capable of developing an antibody response to SARS-CoV-2.
- Younger children were just as likely to be infected as older children and developed similar antibody responses.

## Data Availability

All of the individual participant data collected during this study will be available including data dictionaries on the Queens University Belfast database within 3 months of completion of the study

https://pure.qub.ac.uk/

## Declarations

### • Ethical approval

was obtained from the London – Chelsea Research Ethics Committee (REC Reference – 20/HRA/1731) and the Belfast Health & Social Care Trust Research Governance (Reference 19147TW-SW).

### • Authors contributions

Dr Waterfield, Dr Watson, Dr Ladhani and Dr Christie conceived the study idea. Dr Waterfield, Dr Watson, Dr Ladhani, Dr Christie, Dr Moore, Dr Ferris, Dr McGinn, Dr Foster, Dr Evans, Dr Lyttle, Dr Ahmad, Dr Ladhani, Dr Corr, Dr McFetridge, Dr Mitchell and Dr Maney contributed to the design of the study. Dr Waterfield co-ordinated the running of the study including data management and site training. Dr Corr wrote the study protocol. Dr Lyttle designed the electronic CRFs. Dr Moore co-ordinated and led the PPI group. Dr Christie, Dr Ferris, Dr Foster, Dr Evans, Dr Ahmad and Dr Ladhani were site leads. Dr Tonry, Dr Watson, Dr Amirthalingam, Dr Brown and Dr Watt were responsible for performing laboratory testing. Dr McFetridge and Dr Mitchell provided statistical expertise and performed the statistical analysis. All authors contributed to the writing of the manuscript.

## • Acknowledgements

We thank all of the children and their families who participated in this study. We also thank all of the sites (Belfast Health and Social Care Trust, The Ulster Independent Clinic, Cardiff and Vale University Health Board, NHS Greater Glasgow and Clyde, Public Health England, London, Manchester University NHS Foundation Trust, NIHR Manchester Clinical Research Facility) and staff who participated in screening and enrolment. We also thank St Jude’s Children’s Cancer Aid and Research Institute for providing artwork for the participant information sheet. Individual thanks to Elizabeth Waxman, Derek Fairley, Gala Roew-Setz, James McKenna and Aleksandra Metryka.

## • Data Sharing

All of the individual participant data collected during this study will be available (including data dictionaries) on the Queen’s University Belfast database within 3 months of completion of the study.

## Supplementary Material

STROBE Statement—checklist of items that should be included in reports of observational studies

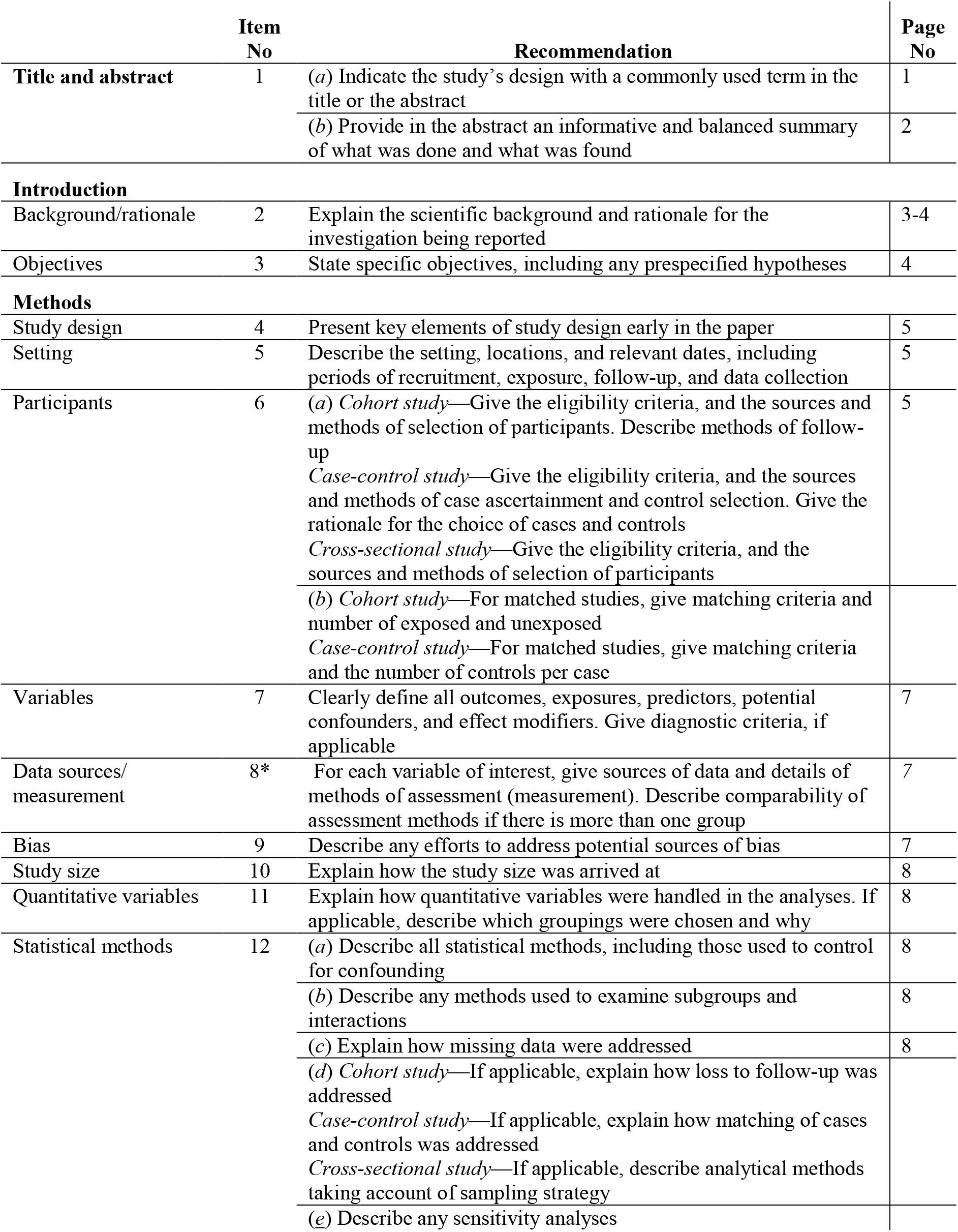
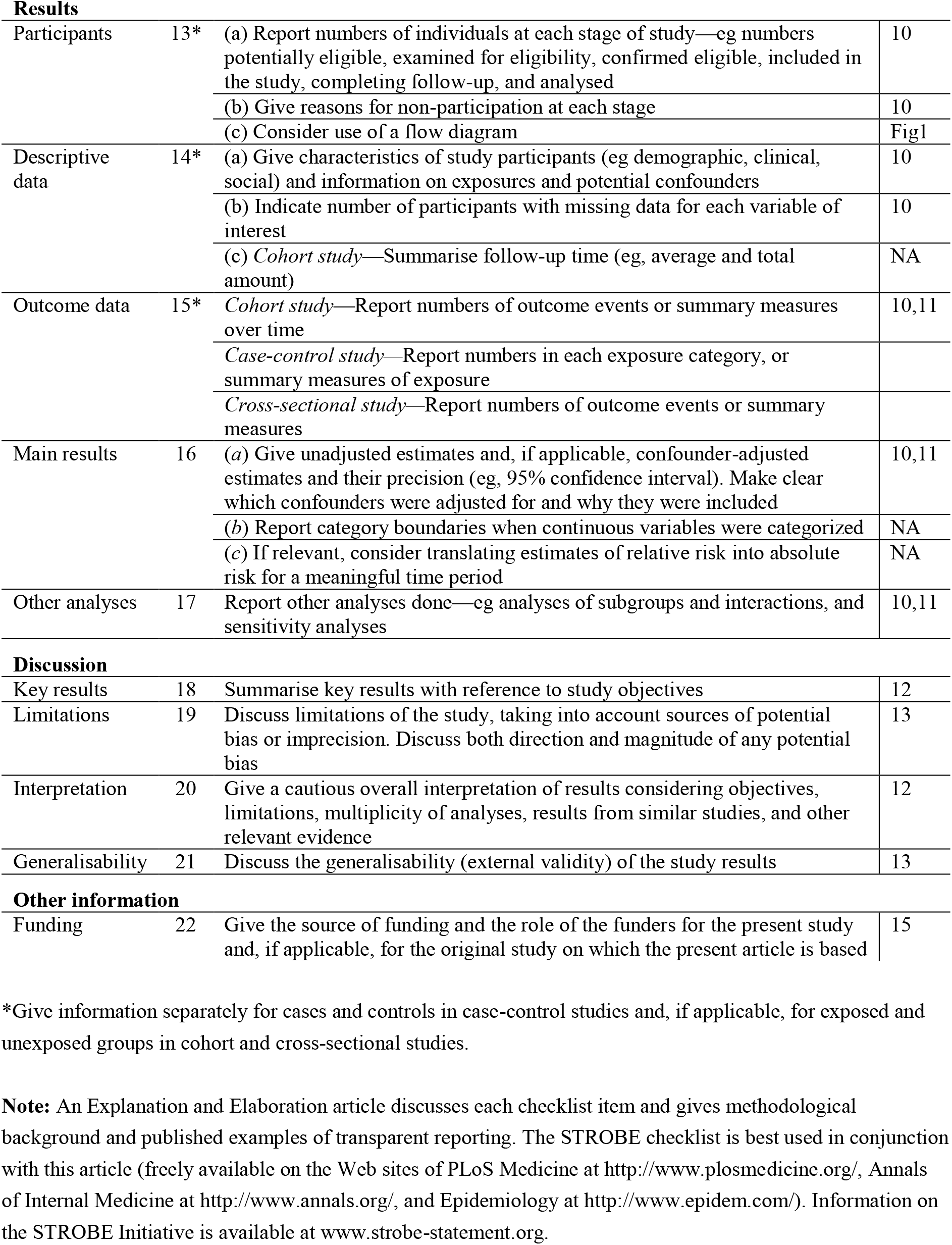

|  | Item No | Recommendation | Page No |
| --- | --- | --- | --- |
| Title and abstract | 1 | (a) Indicate the study’s design with a commonly used term in the title or the abstract | 1 |
|  |  | (b) Provide in the abstract an informative and balanced summary of what was done and what was found | 2 |
| Introduction |  |  |  |
| Background/rationale | 2 | Explain the scientific background and rationale for the investigation being reported | 3-4 |
| Objectives | 3 | State specific objectives, including any prespecified hypotheses | 4 |
| Methods |  |  |  |
| Study design | 4 | Present key elements of study design early in the paper | 5 |
| Setting | 5 | Describe the setting, locations, and relevant dates, including periods of recruitment, exposure, follow-up, and data collection | 5 |
| Participants | 6 | (a) Cohort study—Give the eligibility criteria, and the sources and methods of selection of participants. Describe methods of follow-up<br>Case-control study—Give the eligibility criteria, and the sources and methods of case ascertainment and control selection. Give the rationale for the choice of cases and controls<br>Cross-sectional study—Give the eligibility criteria, and the sources and methods of selection of participants | 5 |
|  |  | (b) Cohort study—For matched studies, give matching criteria and number of exposed and unexposed<br>Case-control study—For matched studies, give matching criteria and the number of controls per case |  |
| Variables | 7 | Clearly define all outcomes, exposures, predictors, potential confounders, and effect modifiers. Give diagnostic criteria, if applicable | 7 |
| Data sources/measurement | 8* | For each variable of interest, give sources of data and details of methods of assessment (measurement). Describe comparability of assessment methods if there is more than one group | 7 |
| Bias | 9 | Describe any efforts to address potential sources of bias | 7 |
| Study size | 10 | Explain how the study size was arrived at | 8 |
| Quantitative variables | 11 | Explain how quantitative variables were handled in the analyses. If applicable, describe which groupings were chosen and why | 8 |
| Statistical methods | 12 | (a) Describe all statistical methods, including those used to control for confounding | 8 |
|  |  | (b) Describe any methods used to examine subgroups and interactions | 8 |
|  |  | (c) Explain how missing data were addressed | 8 |
|  |  | (d) Cohort study—If applicable, explain how loss to follow-up was addressed<br>Case-control study—If applicable, explain how matching of cases and controls was addressed<br>Cross-sectional study—If applicable, describe analytical methods taking account of sampling strategy |  |
|  |  | (e) Describe any sensitivity analyses |  |
| **Results** |  |  |  |
| Participants | 13* | (a) Report numbers of individuals at each stage of study—eg numbers potentially eligible, examined for eligibility, confirmed eligible, included in the study, completing follow-up, and analysed | 10 |
|  |  | (b) Give reasons for non-participation at each stage | 10 |
|  |  | (c) Consider use of a flow diagram | Fig1 |
| Descriptive data | 14* | (a) Give characteristics of study participants (eg demographic, clinical, social) and information on exposures and potential confounders | 10 |
|  |  | (b) Indicate number of participants with missing data for each variable of interest | 10 |
|  |  | (c) *Cohort study*—Summarise follow-up time (eg, average and total amount) | NA |
| Outcome data | 15* | *Cohort study*—Report numbers of outcome events or summary measures over time | 10,11 |
|  |  | *Case-control study*—Report numbers in each exposure category, or summary measures of exposure |  |
|  |  | *Cross-sectional study*—Report numbers of outcome events or summary measures |  |
| Main results | 16 | (a) Give unadjusted estimates and, if applicable, confounder-adjusted estimates and their precision (eg, 95% confidence interval). Make clear which confounders were adjusted for and why they were included | 10,11 |
|  |  | (b) Report category boundaries when continuous variables were categorized | NA |
|  |  | (c) If relevant, consider translating estimates of relative risk into absolute risk for a meaningful time period | NA |
| Other analyses | 17 | Report other analyses done—eg analyses of subgroups and interactions, and sensitivity analyses | 10,11 |
| **Discussion** |  |  |  |
| Key results | 18 | Summarise key results with reference to study objectives | 12 |
| Limitations | 19 | Discuss limitations of the study, taking into account sources of potential bias or imprecision. Discuss both direction and magnitude of any potential bias | 13 |
| Interpretation | 20 | Give a cautious overall interpretation of results considering objectives, limitations, multiplicity of analyses, results from similar studies, and other relevant evidence | 12 |
| Generalisability | 21 | Discuss the generalisability (external validity) of the study results | 13 |
| **Other information** |  |  |  |
| Funding | 22 | Give the source of funding and the role of the funders for the present study and, if applicable, for the original study on which the present article is based | 15 |

\*Give information separately for cases and controls in case-control studies and, if applicable, for exposed and unexposed groups in cohort and cross-sectional studies.
**Note:** An Explanation and Elaboration article discusses each checklist item and gives methodological background and published examples of transparent reporting. The STROBE checklist is best used in conjunction with this article (freely available on the Web sites of PLoS Medicine at http://www.plosmedicine.org/, Annals of Internal Medicine at http://www.annals.org/, and Epidemiology at http://www.epidem.com/). Information on the STROBE Initiative is available at www.strobe-statement.org.

